# Tracheal aspirate metagenomics reveals association of antibiotic resistance with non-pulmonary sepsis mortality

**DOI:** 10.1101/2024.04.08.24305484

**Authors:** Héctor Rodríguez-Pérez, Laura Ciuffreda, Tamara Hernández-Beeftink, Beatriz Guillen-Guio, David Domínguez, Almudena Corrales, Elena Espinosa, Julia Alcoba-Florez, Jose M. Lorenzo-Salazar, Rafaela González-Montelongo, Jesús Villar, Carlos Flores

## Abstract

**Background:** Previous metabarcoding studies based on 16S rRNA sequencing in patients with extrapulmonary sepsis have found early pulmonary dysbiosis associated with a poor prognosis. To further discern this association, here we aimed to better characterize the pulmonary bacterial communities in these patients by leveraging metagenomics and to evaluate if the presence of antibiotic resistance genes (ARGs) could explain the higher mortality of the patients.

**Material and methods:** Metagenomic sequencing was performed using the Nextera XT Library Prep Kit and HiSeq 4000 (Illumina Inc.) on tracheal aspirate samples that were obtained within 24 h from diagnosis from patients with extrapulmonary sepsis admitted to the Intensive Care Unit (ICU). Analysis involved MetaSpades for contig assembly, Kraken2 and Metaphlan4 for taxonomic classification, and CARD and GTDB-tk for ARGs annotation and assignment to the bacterial species. The relationship between the presence of antibiotic resistance and ICU mortality was evaluated using the Wilcoxon test and logistic regression models adjusting for clinical and demographic variables.

**Results:** In total, 127 different ARGs were detected circumscribed only to seven patients. The most common ARGs found were from the antibiotic groups of aminoglycosides and beta-lactams, both present in most of the patients. These ARGs were found, almost entirely linked to *Klebsiella pneumoniae, Escherichia coli*, and *Pseudomonas aeruginosa*. The results also show a significant enrichment of ARGs among patients who died while admitted in the ICU (57%, 95% confidence interval [CI]: 18-90%) compared to surviving patients (20%, 95% CI: 7-40%) (*p*=0.022). Analyses adjusting for clinical and demographic variables did not alter this result.

**Conclusion:** Metagenomic sequencing has allowed an unprecedented characterization of the sepsis lung microbiome showing that antibiotic resistance is common among these patients. The results also suggest a relationship between the early accumulation of ARGs in the lung of patients with extrapulmonary sepsis who die while admitted in the ICU. Studies in independent samples will be needed to validate our findings.

## Introduction

Sepsis is a severe and life-threatening complication of infection characterized by a systemic inflammatory response and organ dysfunction (Singer et al., 2016). Despite significant advances in medical care, sepsis leads the causes of mortality in intensive care units (ICUs) and remains a major cause of morbidity among survivors (Herrán-Monge et al., 2019). The pathogenesis of sepsis is complex and multifactorial, involving interactions between host immune responses and the microbiome (Akrami & Sweeney, 2018; van der Poll et al., 2021).

Rapid sepsis diagnosis and identification of the causative pathogen is crucial for the optimal management of the disease, including the selection of appropriate antimicrobial therapy (Kumar et al., 2006). However, the traditional microbiological culture-based approach for pathogen identification and characterization has several limitations, including the low sensitivity and specificity, the requirement of a pure culture isolation, and the inability to detect fastidious or non-cultivable organisms (Handelsman, 2004). As a result, in >30% of cases the aetiologic pathogen is not identified by these methods (Novosad et al., 2016), reflecting the limitations of current culture-based microbiologic diagnostics.

Consequently, antimicrobial therapies that are initiated early after sepsis diagnosis or ICU admission cannot rely on culture-based information (Strich et al., 2020). However, in this context, the rising trend of antimicrobial resistance (AMR) constitutes a main problem among patients admitted to ICUs, especially for those staying longer periods and under high antibiotic pressure. As a figure, about 0.7 million people die annually worldwide from multidrug-resistant (MDR) strains of microorganisms and it is estimated to increase to 10 million by 2050 (de Kraker et al., 2016).

Our previous studies in mechanically ventilated ICU patients with extrapulmonary sepsis revealed that pulmonary dysbiosis within 8 h from sepsis diagnosis was associated with ICU mortality (Guillen-Guio et al., 2020). These observations offer promising options that could be technologically adapted to rapidly identify those patients at higher risk of death (Rodríguez-Pérez et al., 2022). However, they relied on 16S rRNA sequencing experiments which impedes deep characterization of the bacterial communities that could help to further substantiate the association.

To further discern the association of lung dysbiosis with the ICU mortality among sepsis patients, here we used metagenomic sequencing for improved characterization of the bacterial lung communities and for adding support to the possible enrichment of AMRs among the deceased patients.

## Materials and Methods

### Ethics statement

The study was approved by the Research Ethics Committee of the Hospital Universitario Nuestra Señora de Candelaria (PI-30/14) and performed according to The Code of Ethics of the World Medical Association (Declaration of Helsinki). An informed consent was obtained from all patients or from their legal representatives.

### Study patients, sample preparation, and shotgun sequencing

Samples were collected and stored at -20 °C until use from tracheal aspirates from 32 mechanically ventilated patients (25 survivors, 7 deceased) admitted to the medical-surgical ICU within the first 8 h from sepsis diagnosis (Singer et al., 2016) between January 2015 and January 2019 and followed up until discharge or death. These were critical patients with a diagnosis of acute abdomen and constitute a subset of those with sufficient DNA material from aspirates whose clinical characteristics have been described elsewhere (Guillen-Guio et al., 2020; Rodríguez-Pérez et al., 2022).

DNA was extracted using the QIAmp UCP Pathogen Mini Kit (Qiagen). Following quantification with a Qubit 3.0 fluorometer using a High Sensitivity DNA Analysis Kit (Thermo Fisher Scientific) and sample normalization, libraries were prepared with the Nextera XT Library Prep Kit (Illumina Inc.). Library sizes and concentrations were assessed on a TapeStation 4200 (Agilent Technologies) and the High Sensitivity DNA Analysis Kit. Sequencing was performed using a HiSeq 4000 System (Illumina Inc.) using 150 base paired-end reads along with 1% of a PhiX control V3 (Illumina Inc.).

### Bioinformatic analysis

We designed and deployed a bioinformatics pipeline as a collection of BASH scripts to analyze the obtained metagenomic sequence data (https://github.com/genomicsITER/metagenomics_suppmaterial). In brief, the pipeline consisted in the following stages. Sequences underwent first a quality control step using Trimmomatic v0.39 (Bolger et al., 2014). Also, as a preprocessing step, a rapid taxonomic assignment of reads was performed with Kraken2 v2.1.3 (Wood et al., 2019) to identify the human reads. All the identified human reads were removed from the analyses using KrakenTools v1.2 (Lu et al., 2022). On the remaining reads, the subsequent analysis steps included using MetaPhlAn4 (Blanco-Míguez et al., 2023) to determine the general composition of each sample. In parallel, microbial reads were assembled de novo using Spades v3.15.5 (Bankevich et al., 2012) with a specific parametrization for metagenomic assembly. The contigs generated from the assembly step were then binned with Metabat2 (Kang et al., 2019) to produce contig sets or bins, each one belonging to a different organism. In order to identify the AMRs, all binned contigs were annotated for ARGs based on the Comprehensive Antibiotic Resistance Database (CARD) 3.2.7 (Alcock et al., 2019). The CARD hits with coverage below 85% were excluded from further analysis to prevent false positives. Additionally, we used GTDB-tk v2.3.0 to classify each bin based on the Genome Database Taxonomy (GTDB) v214.1 (Parks et al., 2022) to link the identified ARGs to their respective taxonomy at the species level. Finally, CheckM (Parks et al., 2015) was used to estimate the genome completeness and contamination.

### Statistical analysis

The association between the presence of ARGs and sepsis mortality in the ICU was tested using logistic regression models using R v4.3.0 (R Foundation for Statistical Computing, 2021). Subsequent logistic regression-based sensitivity analyses were performed to investigate the effect of clinical and demographic variables, controlling for the level of diversity as measured by the Shannon diversity index.

### Data availability

The datasets analysed in the study are available from the corresponding author on reasonable request.

## Results

Sequencing data yield an average of more than 32 million reads per sample (range of 10 to 70 million reads) (see **Supplementary Table S1**). After human DNA sequences removal, between 0.25% and 12.8% of the reads per sample identified as non-human were kept for downstream analyses. At this stage, we observed that a higher proportion of non-human reads in the sample was associated with ICU mortality in the cases (Odds Ratio [OR]: 1.37; 95% Confidence Interval [CI]: 1.03-2.21; *p*=0.035).

After metagenomic assembly and genome classification of the non-human reads, 14 genomes with an estimated completeness of 80%-97% and low contamination were assembled across patient samples. All these genomes were classified as *Escherichia coli, Klebsiella pneumoniae, Pseudomonas aeruginosa, Acinetobacter baumannii, Achromobacter xylosoxidans, Staphylococcus aureus*, and *Haemophilus influenzae*, which correspond to the most abundant taxa detected at read level by MetaPhlAn4 in the samples.

ARGs were detected only in nine patients (127 ARGs in total), four of whom died in the ICU (**Figure 1**). We observed that the capacity to detect AMRs was associated with the proportion of non-human reads that were recovered from the samples (OR: 11.02; 95% CI: 1.94-144.58; *p*=0.023). CARD ontology and bin classifications indicated that the most frequent AMR findings were linked to aminoglycosides and beta-lactam antibiotics. These were identified, almost entirely, in the assembled genomes from *E. coli* (48), *K. pneumoniae* (35), and *P. aeruginosa* (31) (**Figure 2**).

**Figure 1.**
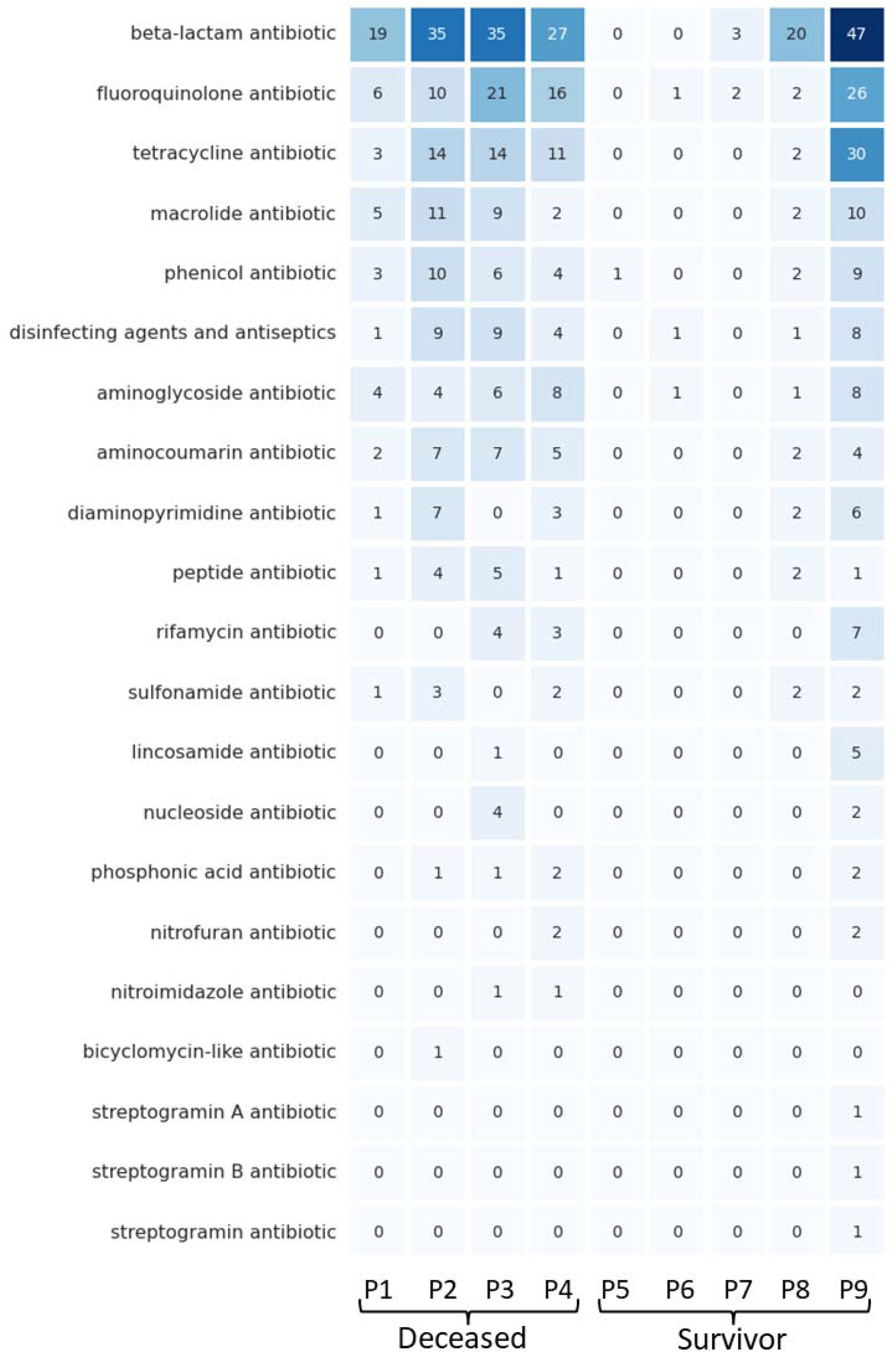
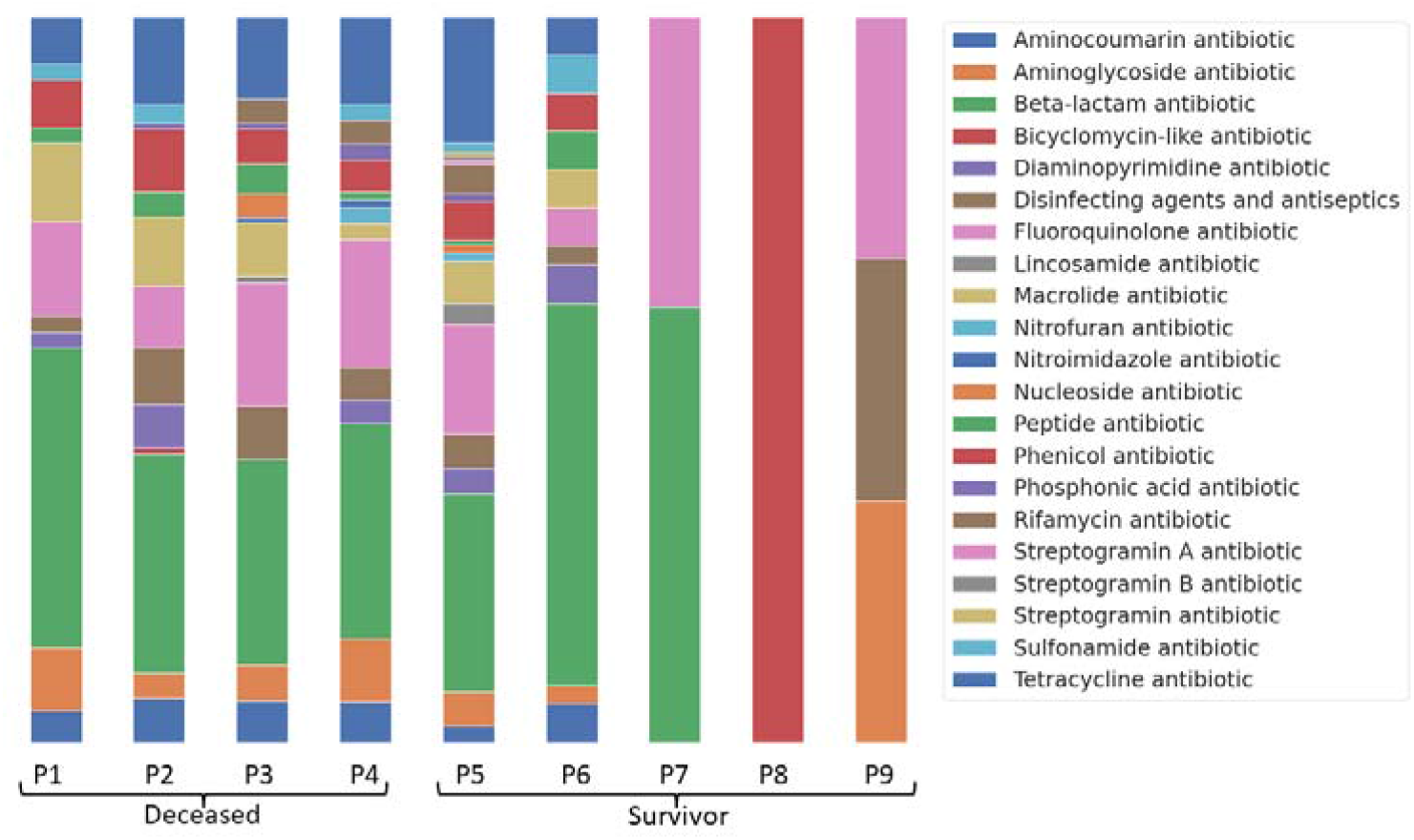
(Upper panel) Absence-presence matrix for AMR resistance groups identified among the nine AMR-positive patients. Numbers in boxes denote the number of different ARGs identified per patient. (Lower panel) Relative abundance of resistances as classified in resistance groups in the nine AMR-positive patients.

**Figure 2.**
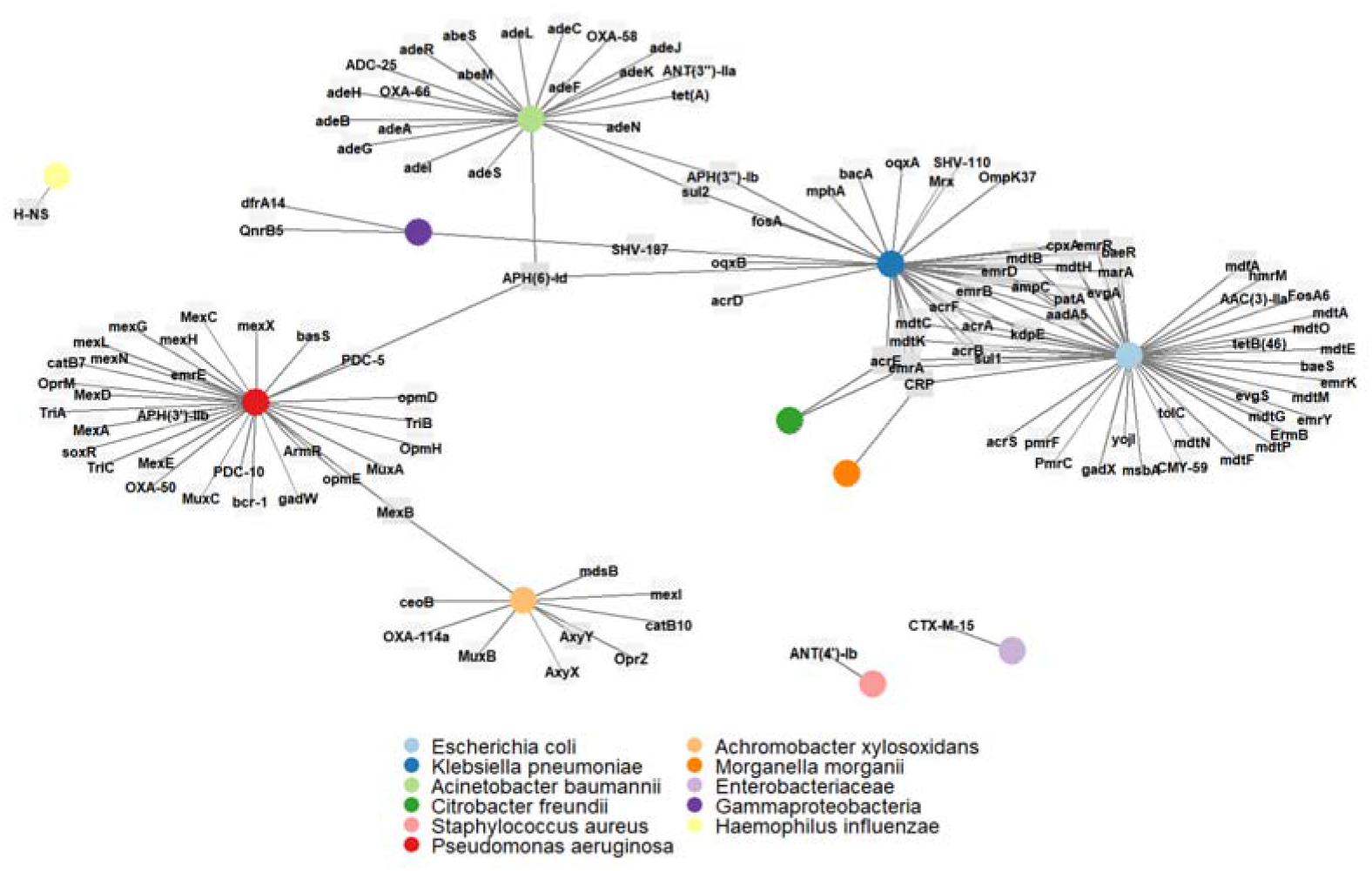
Network depicting the relationships between the AMR resistances and the bacterial taxonomy based on the contigs assembled in the AMR-positive patients.

Finally, we assessed if the abundance of AMRs was associated with ICU mortality in this cohort of patients. We found that the number of AMRs was significantly higher (OR: 1.19; 95% CI: 1.03-1.40; *p*=0.022) in deceased patients (57%, 95% CI: 18-90%) compared to surviving patients (20%, 95% CI: 7-40%). These results were robust to model adjustments for relevant demographic and clinical covariates in the sensitivity analysis (**Table 1**).

**Table 1.** Results from the sensitivity analysis of the model assessing the association of the abundance of AMRs with ICU mortality among extrapulmonary sepsis patients.

| Model adjustment | OR [95% CI] | p-value<sup>a</sup> |
| --- | --- | --- |
| No adjustments | 1.19 [1.03-1.40] | 0.022 |
| *P. aeruginosa* presence<sup>b</sup> | 1.19 [1.02-1.45] | 0.036 |
| Sex | 1.19 [1.03-1.41] | 0.025 |
| Age | 1.18 [1.02-1.40] | 0.035 |
| APACHE II | 1.27 [1.05-1.67] | 0.031 |
| SOFA | 1.30 [1.07-1.70] | 0.017 |
| SAPS II | 1.26 [1.03-1.68] | 0.041 |
| ARDS | 1.30 [1.08-1.69] | 0.012 |
| Smoking | 1.20 [1.04-1.43] | 0.018 |
| Previous severe infections<sup>c</sup> | 1.53 [1.15-3.46] | 0.062 |
| Infection source | 1.18 [1.03-1.40] | 0.028 |
| Isolated pathogen | 1.19 [1.03-1.41] | 0.024 |
| Multiorgan dysfunction<sup>d</sup> | 1.18 [1.02-1.39] | 0.029 |
| Comorbidities<sup>e</sup> | 1.18 [1.02-1.40] | 0.030 |
| Cancer | 1.22 [1.05-1.50] | 0.019 |
| Arterial hypertension | 1.19 [1.03-1.41] | 0.024 |
| Bicarbonate levels (mEq/L) | 1.33 [1.10-1.77] | 0.011 |
| Days in hospital before sepsis diagnosis | 1.47 [1.13-2.33] | 0.026 |
| Days in hospital before ICU admission | 1.19 [1.03-1.41] | 0.020 |

<sup>a</sup>Logistic regression.
<sup>b</sup>Since *P. aeruginosa* typically accumulates a diversity of AMRs, this model adjusted the association for the presence of *P. aeruginosa* in the microbial community. The only two patients where this bacterium was identified had 8 and 28 different ARGs, corresponding to a survivor and a deceased patient, respectively.
<sup>c</sup>According to previous surgical interventions declared in the medical history.
<sup>d</sup>Two or more affected organs based on Sequential Organ Failure Assessment (SOFA) scores.
<sup>e</sup>Presence of comorbidities (personal history): cancer, chronic diseases, diabetes, liver diseases, kidney diseases, previous severe infections, cardiac valve diseases.
APACHE II, Acute Physiology and Chronic Health Evaluation II at inclusion; ARDS, Acute Respiratory Distress Syndrome; CI, Confidence Interval; OR, Odds Ratio; SAPS II, Simplified Acute Physiology Score II; SOFA, Sequential Organ Failure Assessment.

## Discussion

Dysbiosis of the lung microbiome has potential implications for disease pathogenesis and therapeutic interventions (Akrami & Sweeney, 2018; Mukherjee & Hanidziar, 2018). Our previous findings showed reduced lung bacterial diversity at genus and species levels associated with extrapulmonary sepsis mortality (Guillen-Guio et al., 2020; Rodríguez-Pérez et al., 2022). Here, a metagenomics-based analysis allowed us to further characterize the lung dysbiosis in these patients, beyond assessing bacterial composition and diversity, by focusing on genomic assemblies and the identification of AMR profiles. Our analysis revealed that the abundance of AMRs in the lungs within the first hours of sepsis diagnosis correlated with ICU mortality rates, providing a qualitative survey of bacterial co-infections likely contributing to the sepsis severity in these patients and making it the first study linking lung microbiome AMRs to ICU mortality in sepsis patients.

Predominant Gram-negative bacteria like *E. coli, P. aeruginosa*, and *K. pneumonia* harboring diverse resistance mechanisms have been previously identified as common sepsis-causing pathogens (Bloomfield et al., 2023; Mu et al., 2023). Some of these species have been linked to gut-to-lung bacterial migration and AMR acquisition and the worsening of sepsis (Wheatley et al., 2022). A significant proportion of the detected resistance determinants in these species, as well as in most of the AMR-positive samples from our study, belongs to the aminoglycoside, beta-lactam, and peptide antibiotic classes, which have been linked to a substantial impact on the efficacy of antimicrobial treatment (Fatsis-Kavalopoulos et al., 2022; Foudraine et al., 2021). Independent studies have found that poor outcomes in mechanically ventilated critically ill patients were associated with increased bacterial burden in lung microbiome (Dickson et al., 2020). This agrees with our observations where a higher proportion of non-human reads was associated with ICU mortality in our patient series. Taken together, our results emphasize the significance of understanding bacterial pathogens in sepsis severity and early response strategies.

This work has a series of limitations which include a small sample size from a single ICU, preventing optimal confounding effect assessments, and the study’s focus on bacterial reads due to the prevalence of bacterial sepsis, which restricts conclusions regarding non-bacterial pathogens (Faria et al., 2018). In addition, we did not have the possibility to assess an independent sample or access to patient samples before the onset of sepsis to serve as controls. One cannot either conclude that the detected bacteria were pathogens causing the sepsis or if they casually contributed to the patient severity. Moreover, the study relied on a limited proportion of reads non-aligning to the human genome, limiting our ability to assemble highly continuous bacterial genomes which could have consequences in the capacity to improve the detection of AMRs. Despite the advancements in high-throughput sequencing, metagenomic data and antibiotic resistance profiles of lung samples are underrepresented in literature compared to gut or oral microbiomes (Dickson et al., 2016; Ibironke et al., 2020). Within the last decade, metagenomic applications have emerged for precise pathogen identification and gene annotation, albeit challenges like standardized protocols and complex data interpretation remain (Chiang & Dekker, 2020). We suggest that future metagenomic applications could potentially enhance early-response strategies to sepsis, anticipating a growing relevance in clinical applications.

## Conclusion

Sepsis remains a major cause of morbidity and mortality in the ICUs worldwide and early diagnosis and identification of causative pathogens are crucial for optimal patient management. Metagenomics by next-generation sequencing could overcome some of the limitations inherent to the traditional microbiological culture-based approaches and provide important clinical information. Here we applied metagenomics to 32 tracheal aspirate samples from ICU patients with non-pulmonary sepsis to identify the AMRs and to link them to bacterial species. The study results support the association between AMR abundance and patient mortality.

## Data Availability

The datasets analyzed during the current study are available from the corresponding author on reasonable request.

---

**Supplementary Table S1.** Summary of metagenomics sequencing results per patient sample.

| Patient ID | ICU<br>deceased | Total reads | Non-human<br>reads | Non-human<br>reads (%) | Human<br>reads (%) | AMR<br>detected |
| --- | --- | --- | --- | --- | --- | --- |
| P1 | Yes | 36,202,441 | 3,376,440 | 9.327 | 90.673 | Yes |
| P2 | Yes | 41,581,154 | 292,423 | 0.703 | 99.297 | Yes |
| P3 | Yes | 46,674,511 | 764,943 | 1.639 | 98.361 | Yes |
| P4 | Yes | 7,120,230 | 914,503 | 12.844 | 87.156 | Yes |
| P5 | No | 43,891,706 | 3,765,711 | 8.580 | 91.420 | Yes |
| P6 | No | 89,724,869 | 413,662 | 0.461 | 99.539 | Yes |
| P7 | No | 47,351,495 | 219,754 | 0.464 | 99.536 | Yes |
| P8 | No | 23,177,446 | 176,473 | 0.761 | 99.239 | Yes |
| P9 | No | 11,810,435 | 304,998 | 2.582 | 97.418 | Yes |
| P10 | Yes | 23,338,798 | 134,949 | 0.578 | 99.422 | No |
| P11 | Yes | 6,978,421 | 74,547 | 1.068 | 98.932 | No |
| P12 | Yes | 2,189,196 | 46,239 | 2.112 | 97.888 | No |
| P13 | No | 42,139,811 | 180,722 | 0.429 | 99.571 | No |
| P14 | No | 46,216,838 | 175,393 | 0.379 | 99.621 | No |
| P15 | No | 49,424,749 | 155,073 | 0.314 | 99.686 | No |
| P16 | No | 61,660,223 | 157,480 | 0.255 | 99.745 | No |
| P17 | No | 45,026,951 | 115,080 | 0.256 | 99.744 | No |
| P18 | No | 52,359,280 | 167,375 | 0.320 | 99.680 | No |
| P19 | No | 39,310,614 | 84,570 | 0.215 | 99.785 | No |
| P20 | No | 35,944,453 | 246,371 | 0.685 | 99.315 | No |
| P21 | No | 52,792,737 | 169,074 | 0.320 | 99.680 | No |
| P22 | No | 56,261,931 | 138,788 | 0.247 | 99.753 | No |
| P23 | No | 40,066,302 | 484,210 | 1.209 | 98.791 | No |
| P24 | No | 42,011,097 | 126,891 | 0.302 | 99.698 | No |
| P25 | No | 14,426,047 | 84,864 | 0.588 | 99.412 | No |
| P26 | No | 4,703,486 | 110,176 | 2.342 | 97.658 | No |
| P27 | No | 12,427,132 | 150,143 | 1.208 | 98.792 | No |
| P28 | No | 74,324,932 | 1,146,076 | 1.542 | 98.458 | No |
| P29 | No | 2,805,191 | 23,253 | 0.829 | 99.171 | No |
| P30 | No | 10,155,744 | 75,413 | 0.743 | 99.257 | No |
| P31 | No | 8,609,401 | 134,492 | 1.562 | 98.438 | No |
| P32 | No | 10,132,252 | 68,090 | 0.672 | 99.328 | No |
| Mean | - | 33,776,246 | 452,443 | 1.735 | 98.264 | - |

ICU, intensive care unit; AMR, antimicrobial resistance.

